# Population-weighted Image-on-scalar Regression Analyses of Large-Scale Neuroimaging Data

**DOI:** 10.1101/2025.04.21.25326171

**Authors:** Zikai Lin, M. Fiona Molloy, Chandra Sripada, Jian Kang, Yajuan Si

## Abstract

Recent advances in neuroimaging modeling highlight the importance of accounting for subgroup heterogeneity in population-based neuroscience research. When imaging data are available only for a subsample of participants, estimated associations between brain activity and individual characteristics may differ from the corresponding population-average estimand. We develop a population-weighted image-on-scalar regression framework for estimating associations between scalar predictors and high-dimensional neuroimaging outcomes. We apply this framework to functional magnetic resonance imaging (fMRI) data from the Adolescent Brain Cognitive Development (ABCD) Study’s n-back working-memory task, focusing on associations between general cognitive ability and working-memory-related brain activation. The ABCD Study provides baseline weights calibrated to external sociodemographic benchmarks for U.S. children aged 9–10 years; however, the imaging analytic subsample differs from the baseline cohort on several measured child and family characteristics, motivating additional subsample weighting. The proposed approach combines existing baseline population weights with imaging-subsample adjustment weights constructed using inverse propensity score weighting. Simulation studies show that weighted and unweighted estimators perform similarly under correctly specified models, while weighting can improve estimation of population-average coefficient functions when relevant interactions are omitted and selection depends on observed variables. In the ABCD application, weighting changes the magnitude, spatial extent, and statistical detection of brain–cognition association maps, with effects depending on covariate adjustment. Our findings indicate that population weighting adjustments can influence estimated associations between brain activity and cognition and underscore the importance of assessing analytic sample composition and evaluating the sensitivity of results to weighting and model specification.

## 1 Introduction

Neuroimaging research examines how brain activity relates to cognitive, behavioral, social, and environmental characteristics. These brain–behavior associations may vary across individuals and across sociodemographic, environmental, or risk-related contexts. Capturing such variability requires individual-level data from samples that are sufficiently large and diverse to support inference beyond a narrowly defined analytic sample (Dotson and Duarte, 2020). Prior work has shown that sample composition can affect estimated associations between age and structural brain measures, including gray matter volume, cortical thickness, and surface area (LeWinn et al., 2017). Recognizing that small convenience samples often fail to capture this diversity, the field of population neuroscience emphasizes large-scale, population-based designs for evaluating the extent to which neuroimaging findings generalize to broader target populations (Falk et al., 2013; Garavan et al., 2018; Han et al., 2025).

However, generalizability in neuroscience is challenging due to the field’s inherent complexities. Collecting neuroimaging data that accurately reflect a specific target population remains difficult, and most studies rely on convenience samples (e.g., college student volunteers, clinical cohorts), subject to selectivity due to intensive study protocols, limited geographical distributions of study sites, and other logistical barriers. Furthermore, the small effect sizes often require large sample sizes that most studies cannot afford. Data quality may be affected by scanner differences, acquisition protocols, task compliance, and quality-control procedures. Large, multi-site studies with diverse cohorts are therefore essential for distinguishing sample-specific findings from associations intended to describe a broader target population.

The Adolescent Brain Cognitive Development (ABCD) Study was designed to address these limitations by recruiting a large, demographically diverse baseline cohort of U.S. children aged 9–10 (Garavan et al., 2018; Dick et al., 2026). The ABCD Study collected extensive behavioral, cognitive, environmental, and functional magnetic resonance imaging (fMRI) data to characterize child brain development and to examine associations between cognitive and psychological functioning and brain activity (Hagler Jr et al., 2019). Each of the ABCD Study sites has constrained catchment areas, where eligible children must live within reasonable travel distance (e.g., 50 miles) of a major medical center or research facility where fMRI imaging could be performed. Within school districts located in the catchment areas, a probability sample of public and private schools was selected as the basis for the recruitment of the majority of eligible children (3rd and 4th grade students as modal grade levels of 9-10 year olds) to the ABCD baseline cohort. All the children in the targeted age range in selected schools were provided with recruitment materials to take home to their families for participation. The recruitment maintained a national quota of demographic targets based on age, sex, race, and ethnicity (Garavan et al., 2018). However, the process of site selection and obtaining school cooperation and then parental consent could selectively impact the final characteristics of the collected sample (Gard et al., 2023).

Population weights have been developed for the ABCD baseline cohort to calibrate the sample to external sociodemographic benchmarks (Heeringa and Berglund, 2020). These weights are commonly recommended for estimating descriptive population quantities, such as means, proportions, and prevalence estimates. Their role in regression modeling is more nuanced. Weighted and unweighted regression estimates may differ depending on the target estimand, the sampling processes, the variables used to construct the weights, the covariates included in the outcome model, and the relationship between these variables and the outcome or association of interest (Si et al., 2024, 2022; West et al., 2025). Weighting effect on complex, high-dimensional neuroimaging regression models estimates remains unexplored.

This challenge is further compounded when analyzing fMRI task paradigms. Processed imaging data were only available for a smaller subsample compared to the baseline sample, due to non-participation, lack of consent to fMRI data collection, or exclusion during data quality screening with non-compliant to protocols on collection or processing. If inclusion in the analytic imaging sample is associated with measured sociodemographic or contextual characteristics, and those characteristics are also associated with cognitive performance, brain activity, or heterogeneity in brain—cognition associations, then unweighted estimates from the analytic sample may differ from the corresponding population-average estimand (Gard et al., 2023; Rudolph et al., 2023). Weighting can reduce discrepancies between the analytic sample and the target population with respect to measured characteristics, but its validity depends on whether the relevant selection-related variables are measured and adequately incorporated into the weighting procedure. In association studies, weighting adjusts the sample inclusion probabilities so that heterogeneous conditional effects are marginalized over the target population covariate distribution rather than the sample covariate distribution. This corrective approach relies on two core assumptions: (1) *conditional exchangeability* (i.e., selection into the usable imaging subsample is ignorable conditional on observed covariates), and (2) *positivity* (i.e., every subgroup in the target population has a non-zero probability of inclusion).

Image-on-scalar regression models are widely used to estimate associations between scalar predictor variables and spatially distributed, high-dimensional neuroimaging outcomes while accounting for spatial dependencies across brain locations (Zhu et al., 2014; Yu et al., 2021; Zeng et al., 2022). Although prior work has demonstrated subgroup heterogeneity in these spatial brain–behavior mappings across sociodemographic profiles (Lin et al., 2024), most applications of image-on-scalar regression are implemented as if the analytic sample were a simple random sample or as if the sample selection process were ignorable for the target estimand. Consequently, it remains unknown how survey weighting adjustments interact with high-dimensional spatial regression estimators, or whether incorporating weights changes substantive conclusions about brain–cognition associations.

In this study, we evaluate the influence of population weighting on image-on-scalar regression models linking working-memory-related brain activity to cognitive performance in the ABCD Study. Working memory is a foundational cognitive domain with well-characterized neural correlates and developmental significance (Christophel et al., 2017). We define the target population as U.S. children aged 9–10 years corresponding to the external sociodemographic benchmarks used in the ABCD weighting procedure. Our primary estimand is the population-average association between cognitive performance and working-memory-related fMRI activation, represented by the coefficient function in an image-on-scalar regression model. Our objective is to evaluate whether and how incorporating baseline population weights and imaging-subsample selection adjustments changes estimated population-average coefficient maps. We also examine whether the influence of weighting depends on covariate adjustment in the outcome model, thereby clarifying when weighting may alter substantive conclusions in population-based developmental neuroimaging.

## 2 Method

We constructed weights for the imaging analytic subsample in two stages. First, we used the ABCD baseline population weights to account for the baseline sampling procedure. Second, we constructed imaging-subsample adjustment weights using inverse propensity score weighting based on selected baseline characteristics, treating availability of usable working-memory fMRI data as the inclusion indicator (Si et al., 2023, 2024). The product of the baseline population weight and the imaging-subsample adjustment weight was used as the final analysis weight for population-weighted analyses of the imaging subsample. The final weights were normalized to sum to the imaging analytic sample size.

We then incorporated these weights into image-on-scalar regression models to evaluate how weighting affects estimated brain–cognition association maps. To assess the role of model specification, we compared weighted and unweighted versions of models with and without prespecified covariate adjustment. This comparative strategy allowed us to examine whether the influence of weighting depends on the inclusion of covariates in the brain outcome model.

We evaluated the proposed weighted image-on-scalar regression approach using simulation studies and applied it to the ABCD working-memory fMRI data. In the application, we compared weighted and unweighted coefficient maps, inference results, and regions identified as associated with cognitive performance. In this section, we describe the ABCD data and measures, construction of the imaging-subsample weights, weighted image-on-scalar regression estimation and inference, and simulation design.

### 2.1 Data and Measures

The ABCD Study aims to characterize trajectories of child neurodevelopment and the factors influencing youth mental health and cognitive outcomes. Our analysis focuses on associations between working-memory-related brain activity and cognitive ability, which may vary across demographic, social, and environmental contexts (Casey et al., 2018; Sternberg and Grigorenko, 2002).

The imaging outcome variables were cortical-surface contrast maps from the n-back working-memory fMRI task in ABCD Curated Annual Release 2.0.1. Specifically, we analyzed the 2-back versus 0-back contrast. Later ABCD releases contain updated n-back task variables, corrected contrasts, and revised quality-control flags. Therefore, the empirical ABCD application should be interpreted primarily as a methodological illustration of weighted image-on-scalar regression, rather than as a definitive substantive analysis of working-memory activation in the ABCD Study.

The exposure variable of interest was the general cognition measure, the *g*-factor, a composite score derived using confirmatory factor analysis of participants’ behavioral performance on six cognitive tasks: *Picture Vocabulary, Oral Reading Recognition, Flanker, Pattern Comparison Processing, Picture Sequence Memory*, and *Rey Auditory Verbal Learning* (Akshoomoff et al., 2013). These tasks were administered outside the fMRI environment during an in-person assessment (Thompson et al., 2019). The *g*-factor captures general cognitive ability and has been linked to neural activity during cognitive tasks, including working memory (Waiter et al., 2009; Gray et al., 2003). Examining the association between the *g*-factor and the 2-back versus 0-back contrast may identify cortical regions in which working-memory-related activation varies with general cognitive ability (Thompson et al., 2019; Lin et al., 2024).

Control variables in the outcome model included sex, race, ethnicity, household size, parental marital status, household income, and study site. These variables were selected because they may be related to recruitment, imaging-data availability, cognitive performance, or brain activity.

### 2.2 Descriptive Summaries and Subsample Weights

Table 1 compares sociodemographic distributions between the ABCD baseline sample and the imaging analytic subsample used in our analysis. We restricted the baseline sample to complete cases for the variables included in the weighting model, resulting in *n* = 9,905 baseline participants. The imaging analytic subsample included *n* = 3,782 participants with usable working-memory fMRI data.

**Table 1:** Comparison of sociodemographic distributions between the ABCD baseline sample (restricted to complete cases without any missing items) and the imaging subsample (UW: Unweighted, Base-W: Weighted with the base weights, Final-W: Weighted with the final subsample weights). Values are shown as counts (percentages) for unweighted columns and percentages for weighted columns.

| Variable | Baseline sample ($n = 9,905$) | | Imaging subsample ($n = 3,782$) | | |
| --- | --- | --- | --- | --- | --- |
|  | UW | Base-W | UW | Base-W | Final-W |
| **Sex** |  |  |  |  |  |
| Female | 4764 (48.1%) | 49.1% | 1925 (50.9%) | 51.8% | 49.4% |
| Male | 5141 (51.9%) | 50.9% | 1857 (49.1%) | 48.2% | 50.6% |
| **Age** |  |  |  |  |  |
| $\geq 10$ | 4699 (47.4%) | 49.6% | 2064 (54.6%) | 56.8% | 49.8% |
| $< 10$ | 5206 (52.6%) | 50.4% | 1718 (45.4%) | 43.2% | 50.2% |
| **Race/Ethnicity** |  |  |  |  |  |
| Asian | 208 (2.1%) | 3.5% | 74 (2.0%) | 3.4% | 3.6% |
| Black | 1328 (13.4%) | 12.1% | 323 (8.5%) | 7.9% | 11.8% |
| Hispanic | 1853 (18.7%) | 22.0% | 608 (16.1%) | 19.8% | 23.2% |
| Other | 1040 (10.5%) | 6.6% | 362 (9.6%) | 6.0% | 6.8% |
| White | 5476 (55.3%) | 55.8% | 2415 (63.9%) | 62.9% | 54.6% |
| **Parental Marital Status** |  |  |  |  |  |
| No | 2457 (24.8%) | 31.4% | 716 (18.9%) | 25.9% | 31.6% |
| Yes | 7448 (75.2%) | 68.6% | 3066 (81.1%) | 74.1% | 68.4% |
| **Household Income** |  |  |  |  |  |
| $< 50K$ | 2815 (28.4%) | 37.6% | 788 (20.8%) | 30.1% | 37.9% |
| 50–100K | 2834 (28.6%) | 31.5% | 1154 (30.5%) | 34.3% | 31.4% |
| $\geq 100K$ | 4256 (43.0%) | 30.9% | 1840 (48.7%) | 35.6% | 30.7% |
| **Household Size** |  |  |  |  |  |
| $> 3$ | 8254 (83.3%) | 81.7% | 3249 (85.9%) | 83.5% | 80.8% |
| $\leq 3$ | 1651 (16.7%) | 18.3% | 533 (14.1%) | 16.5% | 19.2% |

The ABCD baseline weights use information from the American Community Survey (ACS), a large probability sample survey of U.S. households conducted annually by the U.S. Census Bureau, as external benchmarks for selected demographic and socioeconomic characteristics of U.S. children aged 9 and 10 years (Heeringa and Berglund, 2020). These baseline weights improve alignment between the ABCD baseline cohort and the target population benchmarks and are recommended for population-level descriptive analyses of ABCD data (Gard et al., 2023; Li et al., 2025; Hawes et al., 2025). However, as with any weighting procedure, estimates for small subgroups may remain unstable, and they may not fully account for other factors related to both recruitment and analytic quantities.

As shown in Table 1, comparing to the baseline sample, the imaging subsample includes more female (50.9% vs. 48.1%), 10-year-old (54.6% vs. 47.4%), and White (63.8% vs. 55.3%) children from married families (81.1% vs. 75.2%) with more household members (Household size *>* 3: 85.9% vs. 83.3%) and higher household incomes (Household income ≥100K: 48.7% vs. 43.0%). As a result, the imaging subsample fails to retain participants with low socio-economic status and preserve the same measured sociodemographic composition as the baseline cohort.

Applying the ABCD baseline weights to the imaging subsample reduced some discrepancies but did not fully align the imaging subsample with the weighted baseline cohort. Table 1 shows that, comparing to the weighted ABCD baseline samples, the base-weighted imaging subsample still over-represents 10-year-old (56.8% vs. 49.6%), White (62.9% vs. 55.8%), and children from married families (74.1% vs. 68.6%) with larger household sizes (Household size *>* 3: 83.5% vs. 81.7%) and higher household incomes (Household income ≥100K: 35.6% vs. 30.9%). Applying the baseline weights alone does not account for measured differences associated with availability of usable imaging data.

To mitigate this discrepancy, we constructed imaging-subsample adjustment weights using inverse propensity score weighting. We fit a logistic regression model in the baseline sample with inclusion in the imaging analytic subsample as the binary outcome. The predictors were baseline measures available for both included and excluded participants: age, sex, race, ethnicity, household income, household size, parental marital status, and the *g*-factor. We obtained predicted probabilities of inclusion in the imaging analytic subsample and used their inverse values as imaging-subsample adjustment factors. The final analysis weight was the product of the ABCD baseline population weight and the imaging-subsample adjustment factor.

This two-stage weighting procedure is intended to align the imaging analytic subsample more closely with the target U.S. 9-10-year old population benchmarks on measured characteristics. We selected observed baseline measures related to imaging subsample selection and analytic quantities. It does not guarantee removal of all selection-related differences, particularly if imaging availability depends on unmeasured factors such as detailed motion, task compliance, scanner/protocol features, or processing-specific quality-control indicators that are also related to brain activity or brain–cognition associations.

Table 1 shows that the final weights improved alignment of the imaging subsample with the weighted ABCD baseline sample on the measured variables used in the weighting procedure. Compared with individuals with imaging data, individuals without imaging data had slightly higher baseline weights and substantially higher imaging-subsample adjustment weights, resulting in larger final combined weights for the non-imaging group. Final weights were right-skewed in both groups, with a heavier upper tail among individuals without imaging data, although the coefficients of variation were similar across groups. The baseline and final weight distributions showed substantial overlap between groups, supporting the practical positivity assumption. The more right-skewed adjustment and final weights among individuals without imaging data suggest some areas of weaker overlap, but no extreme weights of concern were observed. As diagnostic checks, we compared weighted and unweighted frequencies of the adjusted factors, as shown in Table 1. The results were robust across alternative weighting models, and truncating the final weights at the 95th percentile produced weighted frequencies very similar to those obtained using the untruncated weights, suggesting that the substantive findings were not driven by limited overlap or by a small number of highly weighted observations. For model fitting, weights were normalized to sum to the analytic sample size.

### 2.3 Weighted Image-on-scalar Regression Model

With the newly constructed final weights, we proceeded to fit the image-on-scalar regression model. To begin, we introduce the model notation. Let 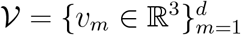 denote the set of *d* vertices in an image, where each vertex *v* corresponds to a three-dimensional spatial coordinate in the image space and is represented as a vector of length 3. For individual *i* = 1, …, *n*, the outcome *y*_*i*_(*V*) ∈ ℝ^*d*^ is collected as a vectorized, spatially distributed image with a set of *d* locations. Let 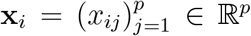 be the *p*-dimensional exposure variables of interest and 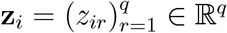 be a vector of *q* control variables.

We model the association between imaging outcome *y*_*i*_(*V*) and scalar variables via the following image-on-scalar regression on the vertex-level:

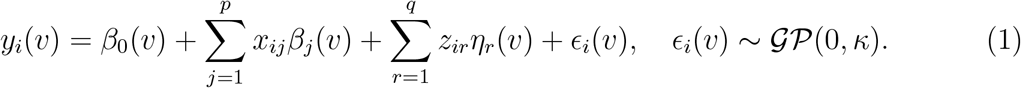

Here *β*_0_(*v*) and 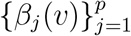 are the spatially-varying intercept function and spatially-varying coefficient (SVC) function of exposure variables 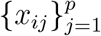, respectively. The spatially-varying coefficient functions 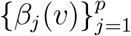 capture the spatial variability of the effect of the exposure variables **x**_*i*_ on the image outcome. The 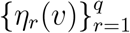 are the SVCs of control variables **z**_*i*_. Since the vertex-space in image data typically exhibits spatial homogeneity, adjacent vertices tend to be correlated, therefore we assume the error term *ϵ*_*i*_(*v*) follows a Gaussian process (*GP*) with zero mean and a kernel covariance function *κ*.

Figure 1 summarizes the model. We are interested in identifying the vertices in the outcome image **y**(*V*) that show significant association with the exposure variable **x** in Model (1). More specifically, we aim to account for the sampling weights and make population-based inferences regarding the coefficients ***β***(*V*).

**Figure 1:**
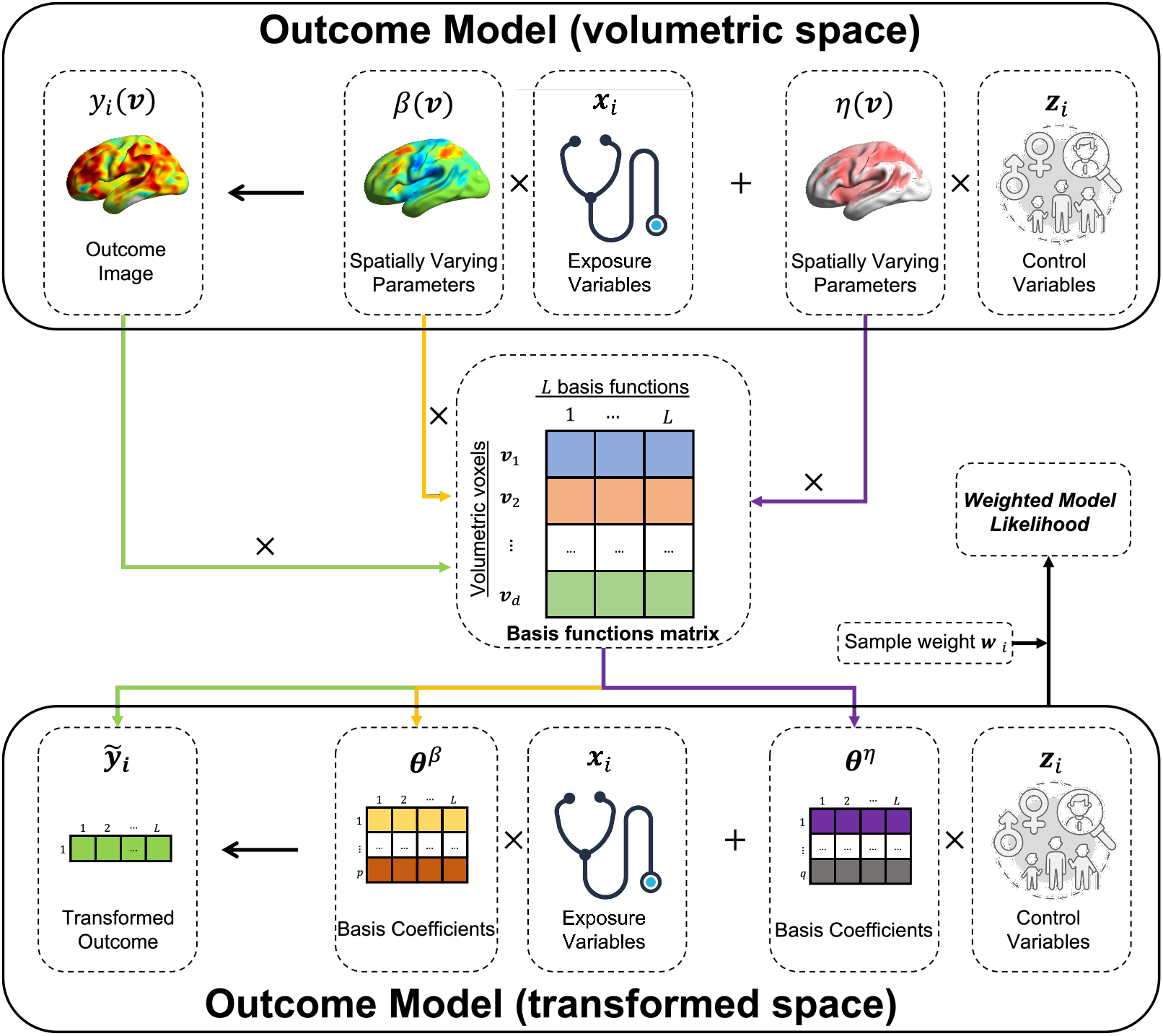
Graphical illustration of the weighted image-on-scalar regression model. The outcome image of individual *i* at vertex *v*, denoted as *y*_*i*_(*v*), is composed of two mean components: 1) The intercept SVCs *β*_0_(*v*) and the association SVCs 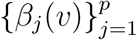 with the exposure variable **x**_*i*_; 2) The association SVCs ***η***_*i*_(*v*) with control variables **z**_*i*_. The volumetric outcome model is converted into a lower-dimensional space by approximating the SVCs using basis function expansions with *L* constant basis functions 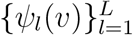 and their corresponding coefficients: 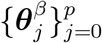 and 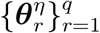. The sample weights *w*_*i*_ are applied to the likelihood functions in the transformed space.

We define 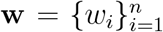 as a vector of the sample weights, with each weight *w*_*i*_ ∈ ℝ^+^ corresponding to the *i*-th observation. In our analysis, we introduce additional notation: ***β***(*V*) represents the set of functions 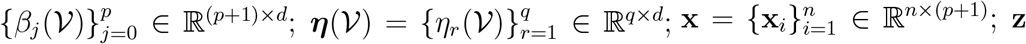 is defined as the set 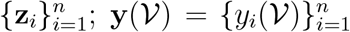; and 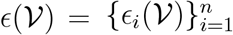. Given the image-on-scalar regression model presented in Model (1), we seek to obtain the estimates of 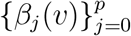 and 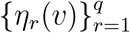 via maximizing the following weighted likelihood function:

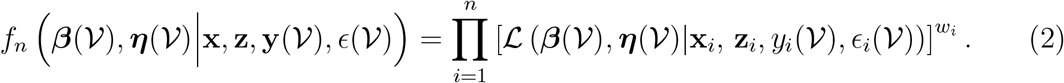

Here, ℒ (***β***(*V*), ***η***(*V*)|**x**_*i*_, **z**_*i*_, *y*_*i*_(*V*), *ϵ*_*i*_(*V*)) denotes the multivariate Gaussian likelihood function.

#### 2.3.1 Model Assumptions

We adopt the basis expansion approach to model the spatial varying coefficients 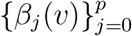 and 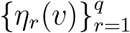 in Model (1). We consider *L* pre-specified basis functions Ψ = *{ψ*_1_, …, *ψ*_*L*_*}* that are orthonormal and satisfy 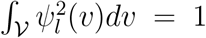 for *l* = 1, …, *L*. We construct the orthonormal basis functions from the eigen-functions of a GP covariance kernel, and the choice of kernel depends on the data structure. For example, we can use the *γ*-exponential kernel function with the form:

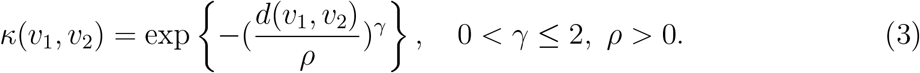

For cortical-surface fMRI data that are mapped onto a spherical surface, *d*(*v*_1_, *v*_2_) denotes the spherical distance between vertices *v*_1_ and *v*_2_, given by *d*(*v*_1_, *v*_2_) = arccos(*v*_1_ · *v*_2_) (Williams and Rasmussen, 2006). The hyperparameter *ρ* controls the smoothness of GP. When *γ* is fixed, larger value of *ρ* indicates a smoother GP kernel. On the other hand, when *ρ* is fixed, a larger value of *γ* indicates a steeper decline in the kernel value with an increasing distance. Note that the *γ*-exponential kernel (3) reduces to the standard squared-exponential kernel when *γ* = 2.

Given the basis functions, we assume the SVCs are approximated by the basis functions expansion:

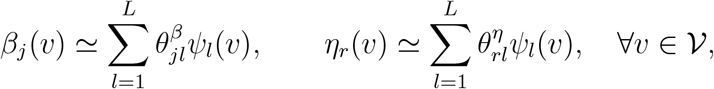

for *j* = 0, …, *p* and *r* = 1, …, *q*. Here, the coefficients 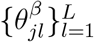 and 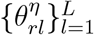 are unknown basis coefficients to be estimated. We also assume that, with the same set of orthonormal basis functions 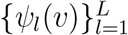, the covariance kernel *κ* defined in (1) can be approximated as:

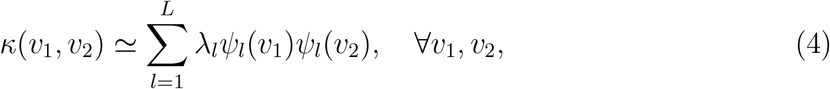

where 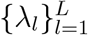 are unknown positive parameters. The positivity constraint of *λ*_*l*_ ensures the positive definite property of covariance kernel *κ* (Ghosal and Van der Vaart, 2017). With the basis functions and basis coefficients defined above, Model (1) becomes:

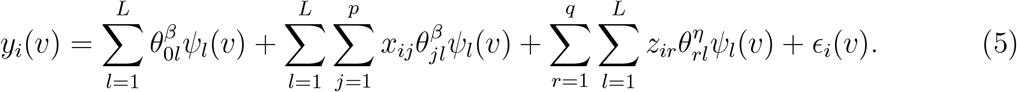

Using a basis expansion approach to represent a SVC reduces the number of parameters from (*p* + *q* + 1) *× d* to (*p* + *q* + 1) *× L*, where *d >> L*, increasing computational efficiency.

#### 2.3.2 Parameter Estimation

We account for the sampling weights and estimate parameters in Models (1), (2) and (5). We first discuss how to obtain orthonormal basis functions 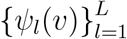 for dimension reduction and then describe the parameter estimation process via maximizing the weighted likelihood function.

We construct 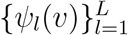 on the set of voxels 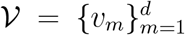 from *L* eigenfunctions, denoted as 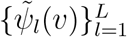 of a pre-specified covariance kernel function on ℝ^3^. First, we evaluate 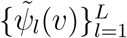 on *V* and obtain the matrix 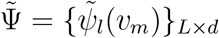. Second we perform the singular value decomposition: 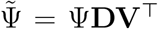. Here, Ψ is a *d × L* rotation matrix of the functional eigenvector and satisfies the orthonormal condition, i.e., Ψ^⊤^Ψ = *I*_*L*_, where *I*_*L*_ is an *L × L* identity matrix. Finally, we use **Ψ** to transform Model (1) into a *L*-dimensional multivariate linear regression model with a diagonal variance structure **Λ** = diag(*λ*_1_, …, *λ*_*L*_). Model (5) can be represented in matrix form as:

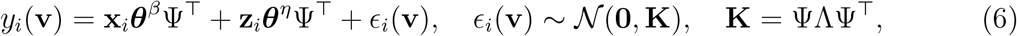

where *ϵ*_*i*_(**v**) = [*ϵ*_*i*_(*v*_1_), …, *ϵ*_*i*_(*v*_*d*_)]^⊤^, and **K** = *{κ*(*v*_*m*_, *v*_*m*_*′*)*}*_*d×d*_. Multiplying both sides of (6) by **Ψ** transforms the model into:

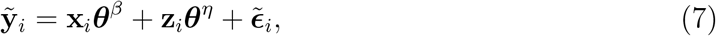

where 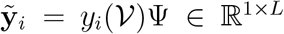 is the transformed response variable, and 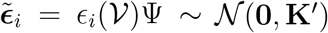 is the transformed error term with **K**^*′*^ = Ψ^⊤^ΨΛΨ^⊤^Ψ = Λ served as the diagonal covariance matrix. Thus, by employing Model (7), we can transform the original *d*-dimensional Model (1) into a more manageable *L*-dimensional multivariate regression problem, where we typically have *d >> L* in real-world applications. Moreover, the transformed random error term 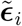 follows a normal distribution with zero mean and a diagonal variance structure Λ, increasing computational efficiency.

We employ maximum likelihood estimation (MLE) to estimate the parameters specified in Model (7). Let **m**_*i*_ = [**x**_*i*_, **z**_*i*_] ∈ ℝ^1*×*(*p*+*q*+1)^ denote the concatenated predictor vector and 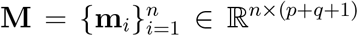 represent the combined design matrix. The combined basis coefficients can be represented by ***θ*** = [***θ***^*β*^, ***θ***^*η*^] ∈ ℝ^(*p*+*q*+1)*×L*^, and the transformed outcome matrix is denoted as 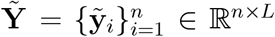. Model (7) can be represented in the matrix form: 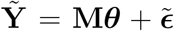. The corresponding weighted likelihood function 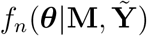 can be defined as:

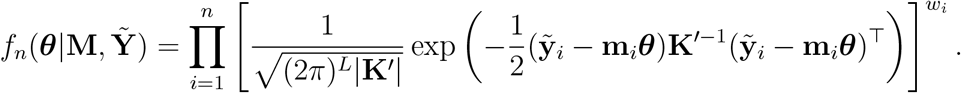

We use **W** = diag(*w*_1_, …, *w*_*n*_) to represent the weight matrix, where each diagonal element corresponds to the weight of an individual. We solve the following score equations and estimate the population parameters ***θ***:

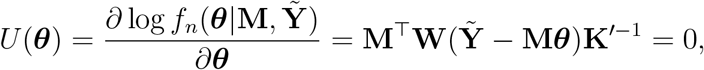

and we obtain:

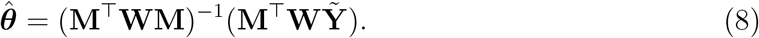

Based on the orthonormal properties of the basis functions matrix **Ψ** discussed above, we can derive the estimates of the SVCs, *{****β***(*V*), ***η***(*V*)*}* through a linear transformation of 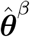 and 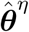:

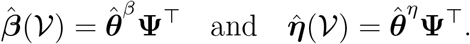

#### 2.3.3 Inference on Population Coefficients

With sample survey data, the variance estimation of coefficient estimates needs to account for the sampling variance introduced by the weights (Rust, 1985). Since the covariance matrix **K**^*′*^ is diagonal, we use the delta-method (Binder, 1983) to obtain the “sandwich” form estimator for the covariance of the coefficient estimates 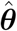

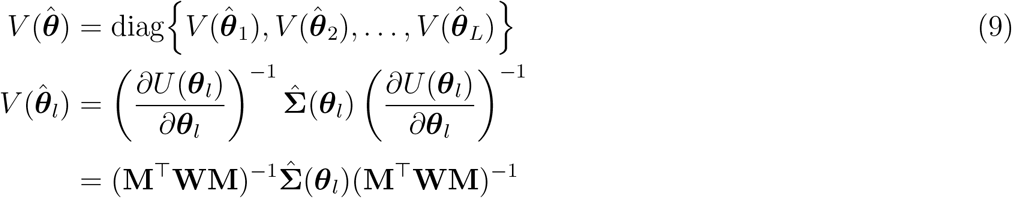

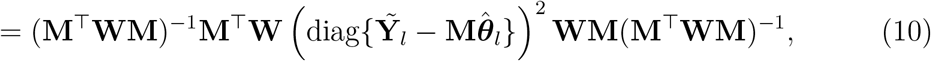

for *l* = 1, …, *L*, where we use the White’s heteroscedasticity-consistent estimator to replace 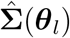 (White, 1980).

The following proposition gives the representation of 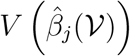.

##### Proposition 2.1.

*Given the variance-covariance matrix* 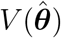 *for* 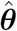, *since the SVC* 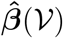 *is a linear transformation of* 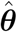, *i*.*e*., 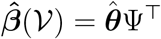. *the covariance matrix for the vectorized* 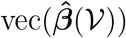 *can be expressed as:*

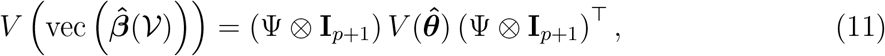

*where* 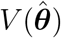 *is defined in* *Equation* (9).

*Proof*. Since ***β***(*V*) = ***θ***Ψ^*T*^, by using the Kronecker product property for the vectorization operator vec(·), we have 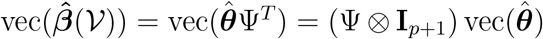. We apply this for the covariance of a Kronecker product:

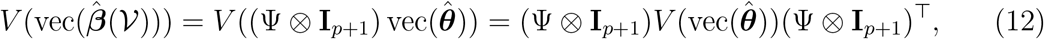

where 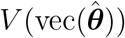 is the estimated variance-covariance matrix of 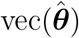. The variance-covariance matrix *V* (vec(***θ***)) is given by

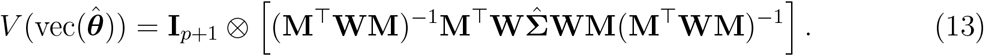

Combining (12) and (13), we have

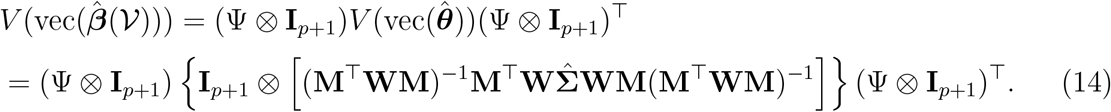

This completes the proof.

The diagonal elements of 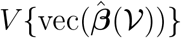 provide the variance estimates of ***β***(*V*) evaluated at each vertex *v* for *j* = 0, …, *p*. With the estimated variance 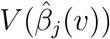, we test the null hypothesis *H*_0_ : *β*_*j*_(*v*) = 0 against *H*_1_ : *β*_*j*_(*v*)≠ 0 using the Wald statistic

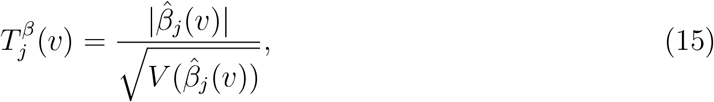

for *j* = 0, …, *p* and *v* ∈ *V*.

After computing the two-sided Wald test statistics at each vertex *v*, we generate a *p*-value image. For each regression coefficient and fitted model, we apply the Holm adjustment across vertices separately within the left and right hemispheres. Vertices with Holm-adjusted *p*-values at or below 0.01 are retained, controlling the family-wise error rate within each coefficient-by-model-by-hemisphere testing family at 0.01. To facilitate interpretation, we summarize vertices with significant associations at the functional-region level using the Gordon 333-region cortical parcellation (Gordon et al., 2016).

The variance estimator treats the constructed weights as fixed. This approach is common in applied weighted regression analyses but does not fully propagate uncertainty from estimating the imaging-subsample propensity weights. We return to this limitation in the Discussion.

### 2.4 Simulation Design

We describe the data generation processes in the simulation studies to examine the impact of including survey weights. We generated a synthetic population with *N*_pop_ =1,000,000 of individuals, from which we repeatedly drew samples and fit image-on-scalar regression models. We mimicked real-life scenarios where it is often impractical to correctly specify a model. We considered a data generation model and two fitting scenarios as the following: 1) a correctly-specified model the same as the data generation model, which includes two-way interaction terms between control variables and the exposure variable, and 2) a mis-specified model that ignores the interaction term.

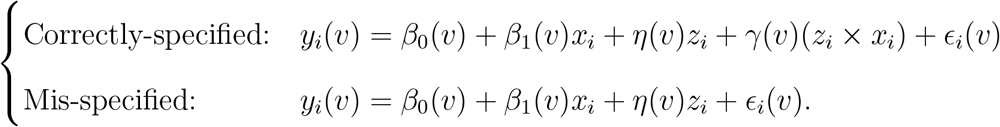

We considered univariate exposure and control variables, both of which were generated from a standard normal distribution, i.e., *x*_*i*_ ~ *N* (0, 1) and *z*_*i*_ ~ *N* (0, 1). We used a logistic model to calculate the inclusion probability Pr(*δ*_*i*_ = 1|*z*_*i*_) for each individual to be sampled, with a logit function, logit(Pr(*δ*_*i*_ = 1|*z*_*i*_)) = −1 − *z*_*i*_. We generated the binary inclusion indicator *δ*_*i*_ for each participant from a Bernoulli distribution: *δ*_*i*_ ~ Bernoulli (Pr(*δ*_*i*_ = 1|*z*_*i*_)). Given the covariates and coefficients, we simulated the outcome image for each subject in the population using the following data generation model (DGM):

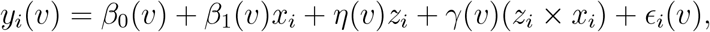

for *v* ∈ *V* and *i* = 1, …, *N*_pop_. The true *β*_0_(*v*) and *β*_1_(*v*) are plotted in the top-row of Figure 2. The parameter *γ*(*v*) represents the SVC for the interaction term (*z*_*i*_ *× x*_*i*_) and is from an approximate Gaussian process as follows:

**Figure 2:**
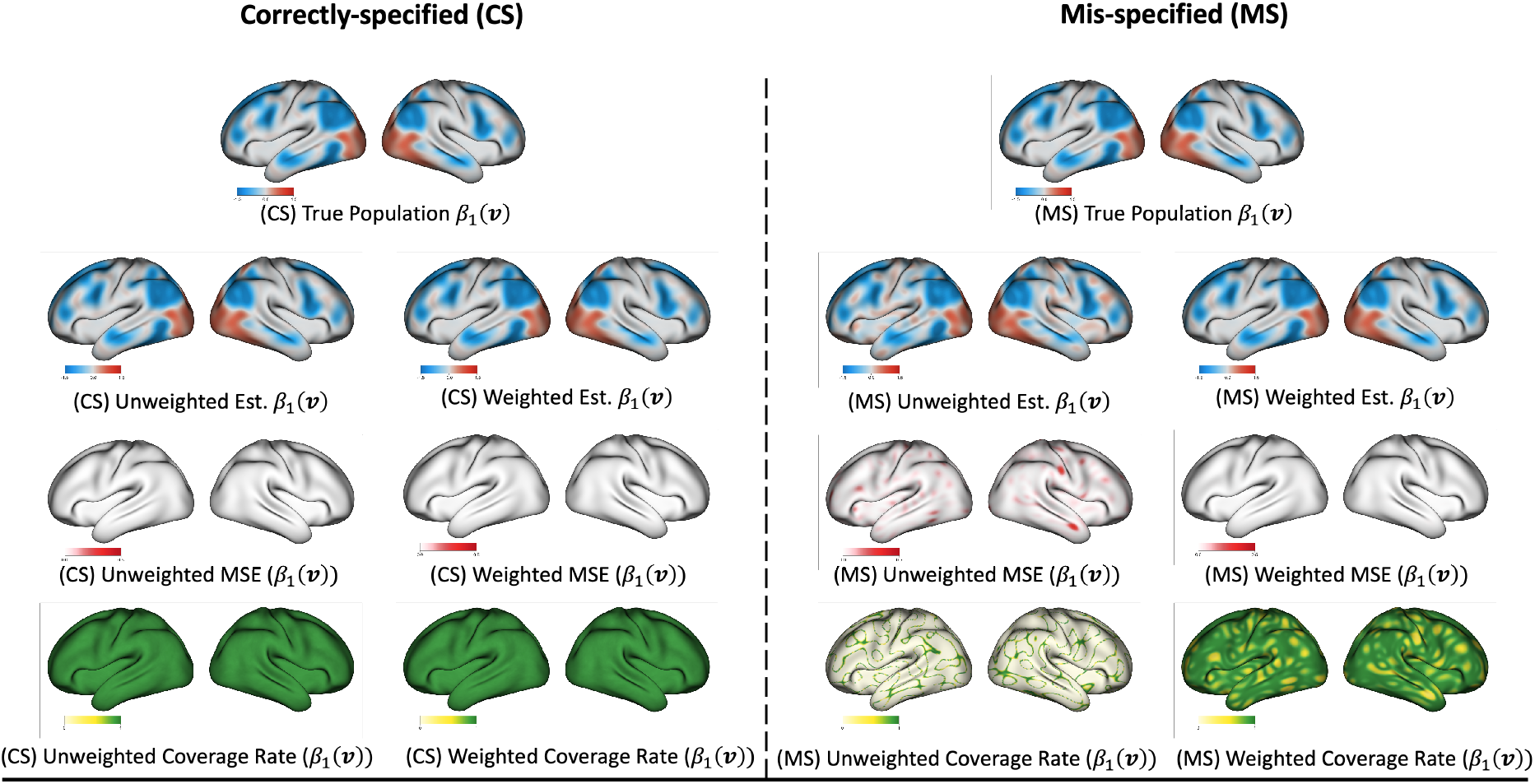
Visual representation of SVC slope *β*_1_(**v**) across different model specification. Top row: true population coefficients of slope SVC *β*_1_(**v**). Second row: point estimation by unweighted and weighted image-on-scalar regression. Third row: average mean squared error across replicates by unweighted and weighted methods. Bottom row: average coverage rate across replicates by unweighted and weighted methods.

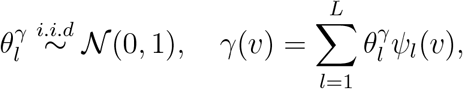

where 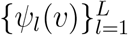 are orthonormal basis functions discussed in Section 2.3.2. In our simulation studies, we set *L* = 500. We generate error terms *ϵ*_*i*_(*v*) from an approximated Gaussian process with zero mean and a covariance function defined as 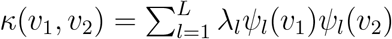. We adjust the signal-to-noise ratio by varying values of pre-specified eigenvalue *λ*_*l*_, each taking on values proportionate to the eigenvalues of the kernel covariance function *κ*.

We repeatedly sampled *n* = 2000 cases from the population using the selection probabilities Pr(*δ*_*i*_ = 1|*z*_*i*_) 500 times. The graphical illustration of the DGM and sample selection is presented in Figure 3.

**Figure 3:**
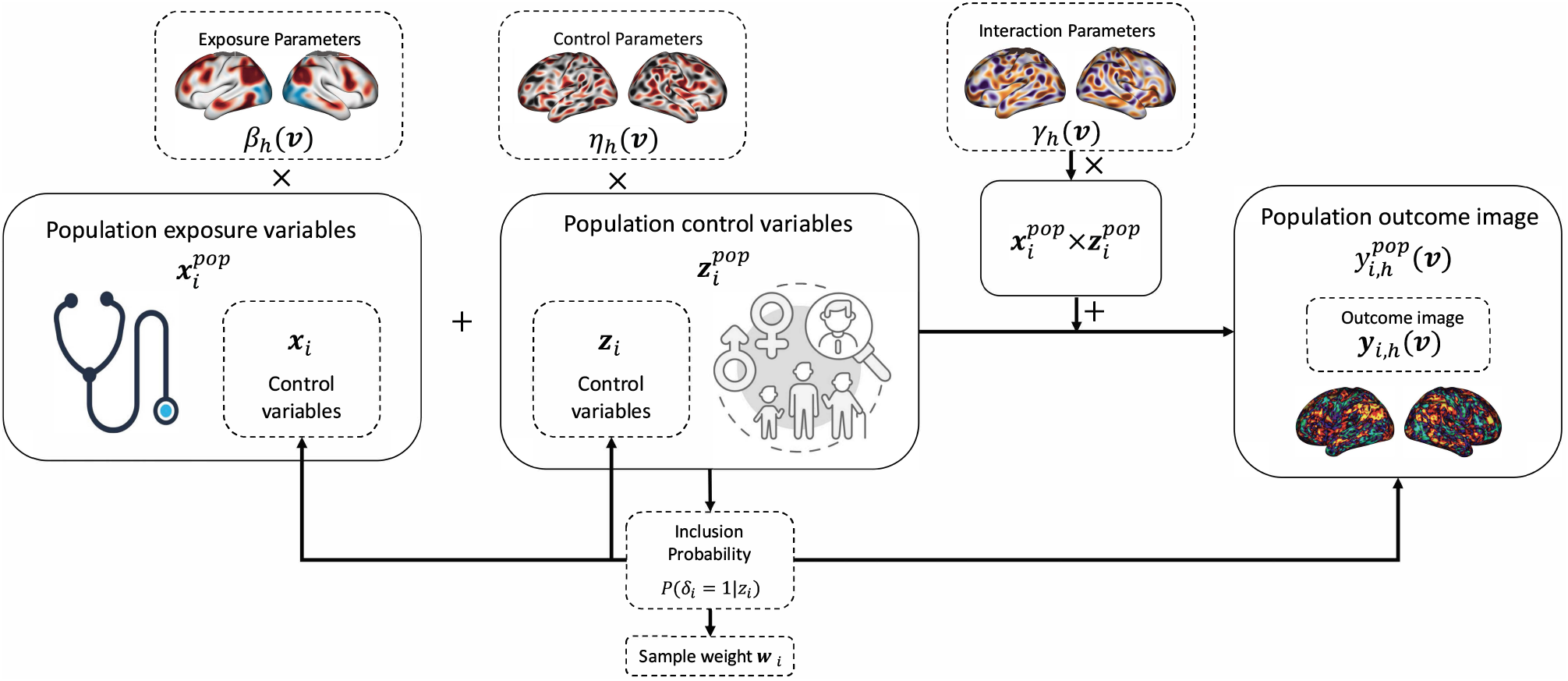
Illustration of the data generation model and sample selection for the simulation study: The population-simulated outcome images, denoted as 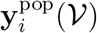, were generated using population exposure variables 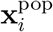 and their associated exposure parameters ***β***(*V*), population control variables 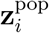 and their associated control parameters ***η***(*V*), along with interaction terms 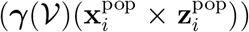. The population control variables 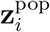 were input into a predetermined logistic regression model to determine the inclusion probabilities for each individual. Within each replicate, subsamples were drawn from the population based on these inclusion probabilities. The sample weights **w** were computed as the inverse of the inclusion probabilities.

We calculated the mean squared error (MSE) of the point estimates *β*_1_(*v*) and averaged their values across all voxels *v* ∈ *V* and all replicates. Additionally, to assess the robustness of the model inferences in both weighted and unweighted methods, the true coefficient coverage rate over 500 sets of the 95% confidence intervals was computed for each voxel *v* ∈ *V*. We reported four primary performance metrics: bias, MSE, coverage rates, and FDR. These metrics were determined across the vertex space for both weighted and unweighted methods, using 500 replicates.

It is important to note that when obtaining the “true coefficient” values in the mis-specified model, we should refer to the coefficients estimated from the finite population. However, with a population size of *N*_pop_ =1,000,000, obtaining the population estimates of *β*_0_(*v*), *β*_1_(*v*), and *η*(*v*) can be computationally demanding. This challenge becomes particularly noticeable when performing regression analysis on the transformed outcome 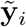 on *x*_*i*_ and *z*_*i*_. To address this computational challenge, we employed a stochastic gradient descent (SGD) method (Diebolt and Ip, 1996) to estimate the finite population coefficients. This approach iteratively updates the estimates of the population-level coefficients using the gradient of the loss function and reduces the computational burden associated with transforming the outcome and estimating the coefficients.

The SGD method for estimating the population coefficients can be summarized as follows. We define the design matrix as 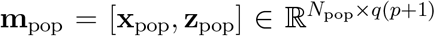. Additionally, we denote the concatenated basis coefficients as 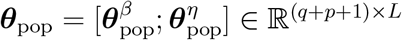. Our goal is to minimize the loss function 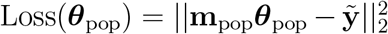. To update the coefficients, we calculate the derivative of the loss function with respect to ***θ***_pop_ as follows:

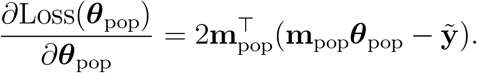

During the process of generating a sample (**m**_*i*_, 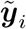) from the population, we update the estimates of the population coefficients 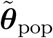 using the gradient descent algorithm:

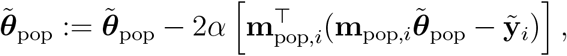

where *α* represents the learning rate of the gradient descent algorithm. This iterative approach updates the population-level coefficients and allows us to estimate ***θ***_pop_ effectively. In comparison to the least square estimator, which has a computation complexity of 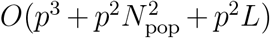 for estimating population-level basis coefficients, SGD reduces the computation complexity to just *O*(*N*_pop_*p*^3^ + *n*_pop_*p*^2^*L*), by circumventing extensive matrix multiplications.

## 3 Results

In this section, we present the results from both the ABCD Study application and simulation studies.

### 3.1 The ABCD Data Analysis Results

We applied the proposed weighted image-on-scalar regression framework to the ABCD working-memory fMRI data. As noted above, the empirical analysis used ABCD Curated Annual Release 2.0.1. Because later releases include updated n-back contrasts, the present application should be interpreted primarily as a methodological illustration of how population and imaging-subsample weights can affect image-on-scalar regression estimates.

We fit weighted and unweighted image-on-scalar regression models, both with and without adjustment for the control variables **z**_*i*_ described in Section 2.1. Our primary focus was inference on the spatially varying coefficient *β*_1_(*V*) for the exposure variable of interest, the *g*-factor. In the weighted analyses, the coefficient function is interpreted as an estimate of the population-average association between the *g*-factor and working-memory-related fMRI activation under the assumptions of the weighting procedure. In contrast, the unweighted analyses estimate the corresponding association in the available imaging analytic sample.

Figure 4 shows the estimated coefficient maps for weighted and unweighted models, with and without covariate adjustment. Across model specifications, both weighted and unweighted analyses identified spatial clusters in which the *g*-factor was positively associated with working-memory-related activation. These clusters were observed primarily in frontoparietal and attention-related regions, including areas within macroscale networks described by Gordon et al. (2016), such as the Dorsal Attention Network (DAN), Fronto-Parietal Network (FPN), and Salience Network (SN). Both approaches also identified regions with negative estimated associations, including vertices in the Cingulo-Opercular Network (CON), Default Mode Network (DMN), and Parieto-Occipital Network (PON).

**Figure 4:**
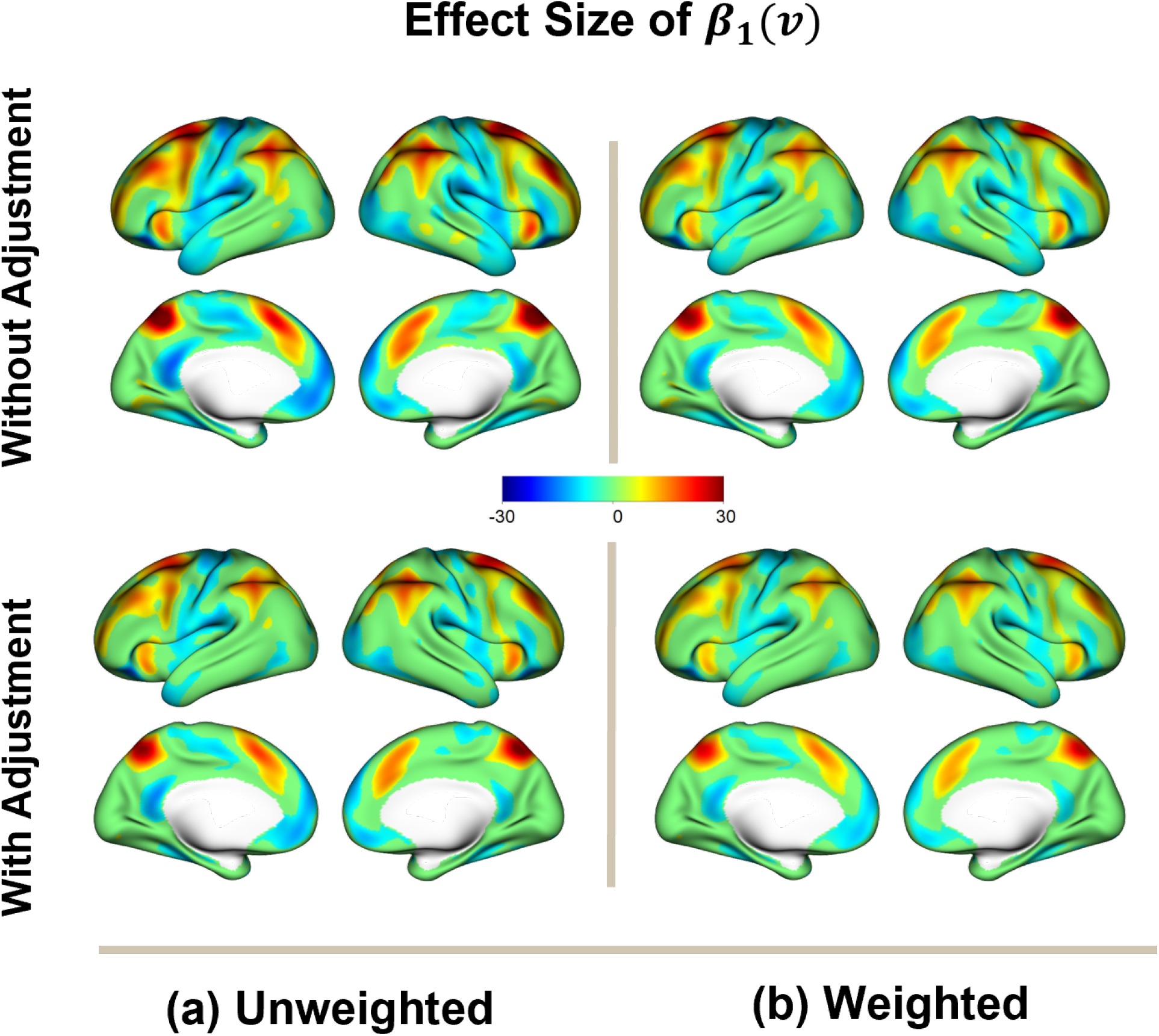
Brain maps of estimated spatially varying coefficients *β*_1_(*V*) from unweighted and weighted image-on-scalar regression models. The top row shows models without adjustment for control variables, and the bottom row shows models with control-variable adjustment. Displayed vertices had Holm-adjusted *p*-values at or below 0.01, with adjustment performed across vertices separately within each hemisphere.

These results indicate that the broad spatial pattern of estimated *g*-factor associations was qualitatively similar across weighted and unweighted analyses. However, the magnitude, spatial extent, and statistical detection of associations differed across weighting and covariate-adjustment specifications. These differences should be interpreted as evidence that population and imaging-subsample weighting can affect estimated coefficient maps in this analytic dataset, rather than as evidence that one map is uniformly correct and the other is biased.

Figure 5 summarizes vertices for which statistical detection differed between weighted and unweighted analyses after applying the Holm adjustment across vertices separately within each hemisphere, using an adjusted *p*-value threshold of 0.01. In this application, the unweighted models generally detected a larger number of significant vertices than the weighted models, with many differences occurring near the boundaries of larger detected clusters. Regions detected only in the unweighted analyses included portions of the insula, middle temporal gyrus, cingulate cortex, primary motor cortex, primary somatosensory-hand cortex, and primary auditory cortex. Conversely, some vertices, including portions of the inferior temporal gyrus, were detected only in the weighted analyses.

**Figure 5:**
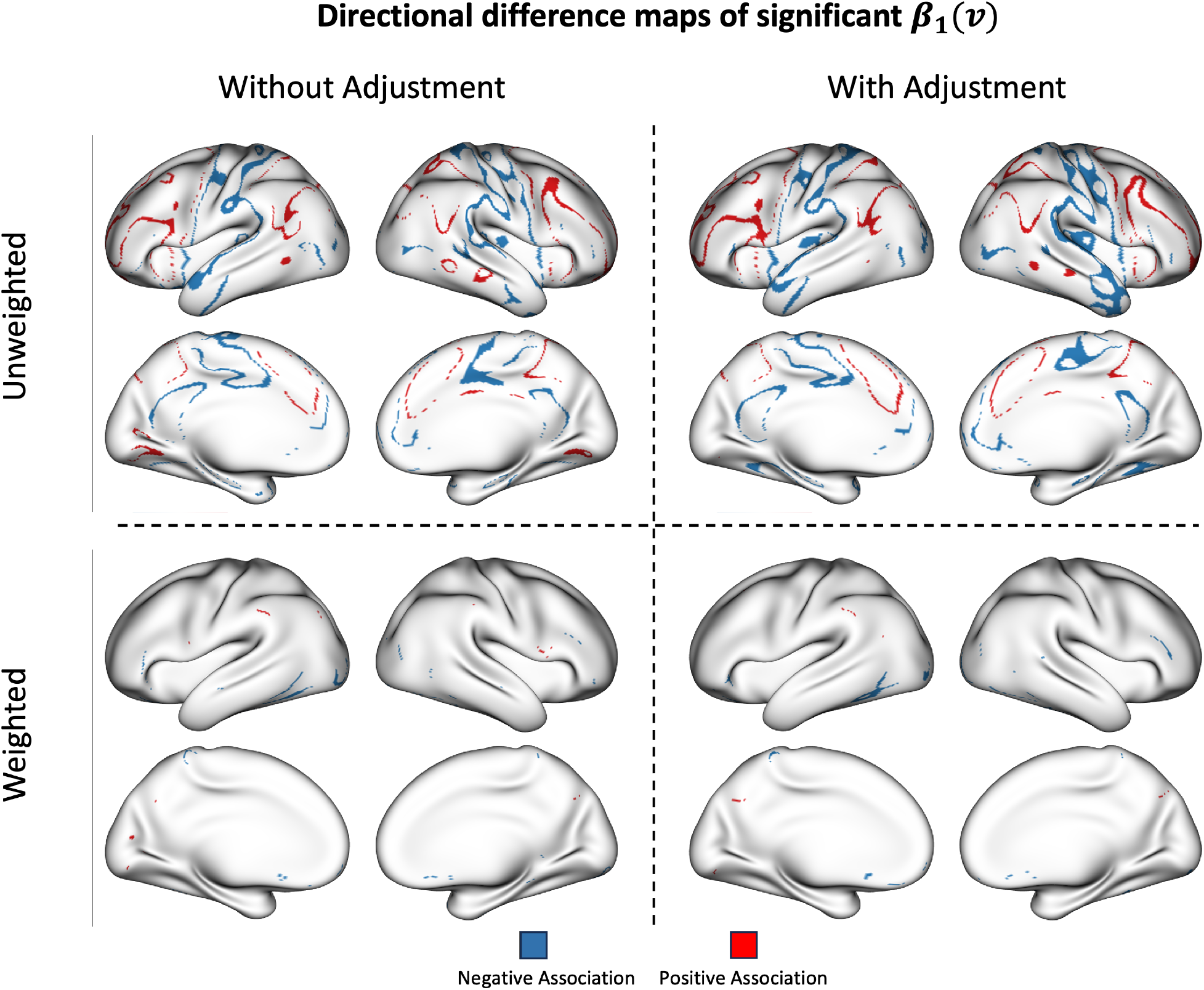
Directional binary maps of vertices differentially detected by weighted and unweighted image-on-scalar regression models for *β*_1_(*v*) after applying the Holm adjustment across vertices separately within each hemisphere, using an adjusted *p*-value threshold of 0.01. Columns show results without control-variable adjustment (left) and with control-variable adjustment (right). The maps distinguish vertices detected only by the unweighted model from vertices detected only by the weighted model.

The comparison also illustrates that the impact of weighting depended on model specification. For example, in the left lateral occipital cortex, differences between weighted and unweighted detection were reduced after adjustment for control variables. In contrast, differences in portions of the cingulate cortex within the frontoparietal system persisted even after covariate adjustment. These patterns are consistent with the expectation that the effect of weighting in regression models depends on the relationship among the weighting variables, the covariates included in the outcome model, and the spatially varying outcome. Because the weights were constructed using measured baseline characteristics and imaging-subsample adjustment variables, these comparisons should be interpreted as sensitivity analyses to weighting choices under measured-variable assumptions.

### 3.2 Evaluation of the Application Study Results

We further evaluated the fitted models using cross-validation within the observed imaging analytic sample. Specifically, we performed 100 random splits of the data. In each replicate, 80% of the analytic sample was used for model fitting and the remaining 20% was held out for prediction. For each cortical vertex, we computed the predictive root mean squared error (RMSE) between the observed and predicted imaging outcomes in the held-out test set. We then compared weighted and unweighted regression models, both with and without control-variable adjustment, by calculating the percentage of vertices within each functional network for which the weighted model had a smaller RMSE than the corresponding unweighted model.

Table 2 reports, for each functional network, the percentage of vertices at which the weighted model produced a lower held-out RMSE than the corresponding unweighted model. In models without control-variable adjustment, the weighted model had lower RMSE for more than half of the vertices in several networks, including the Auditory, Cingulo-Opercular, Default Mode, Dorsal Attention, Ventral Attention, Visual, Fronto-Parietal, and Somatomotor Hand networks. This suggests that, in unadjusted models, incorporating weights did not uniformly reduce predictive performance in the observed analytic sample and in some networks yielded modestly better held-out prediction.

**Table 2:**
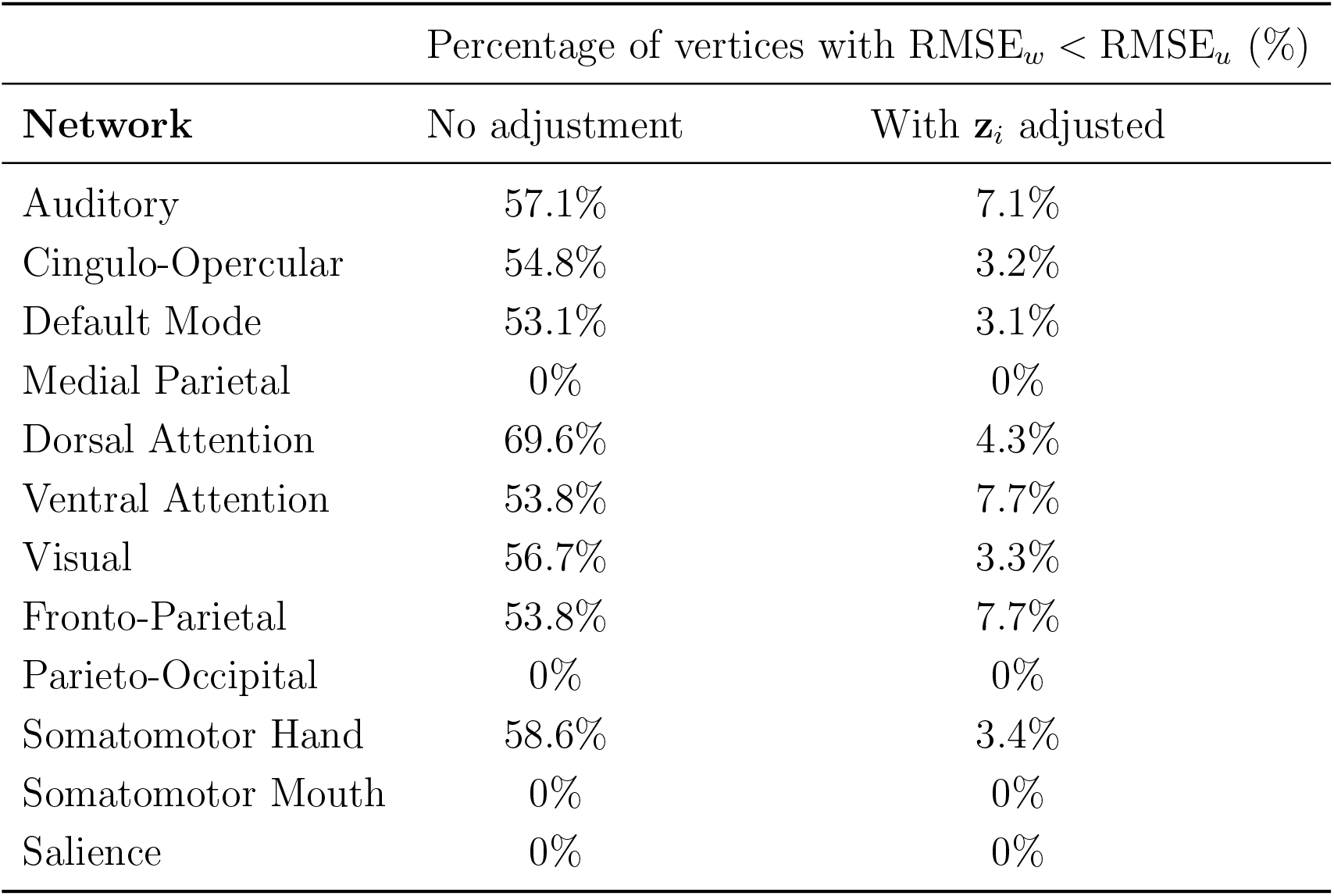
Percentage of vertices within each functional network for which the weighted image-on-scalar regression model had lower predictive RMSE than the corresponding unweighted model. Results are shown for models without and with adjustment for control variables z_*i*_.

In contrast, after adjustment for control variables **z**_*i*_, the unweighted models had lower RMSE for most vertices across all networks. This finding indicates that, within the observed imaging analytic sample, covariate-adjusted unweighted models provided better held-out prediction than the corresponding weighted models. This result should not be interpreted as evidence that the unweighted estimates are necessarily more appropriate for population-average inference. Rather, it highlights that predictive performance in the analytic sample and estimation of a target population-average association are related but distinct goals.

It is important to note that this cross-validation analysis evaluates predictive performance in held-out observations from the available imaging analytic sample. It does not directly validate population generalizability, nor does it determine whether the weighted or unweighted estimator is closer to the target population-average estimand. Weighted and unweighted analyses may optimize different objectives: unweighted models may predict better for the observed analytic sample, whereas weighted models are intended to align estimation with the target population under the assumptions of the weighting procedure.

Together, the coefficient maps, differential-detection maps, and cross-validation results show that the influence of weighting depends on model specification. In particular, weighting had a larger impact in models without covariate adjustment and a smaller, though still spatially heterogeneous, effect in covariate-adjusted models. These findings support the use of weighted and unweighted comparisons as sensitivity analyses when applying high-dimensional neuroimaging regression models to analytic samples that may differ from the intended target population

### 3.3 Simulation Results

We conducted simulation studies to compare weighted and unweighted image-on-scalar regression estimators for the spatially varying coefficient *β*_1_(*v*) under correctly specified and misspecified outcome models. We evaluated bias, mean squared error (MSE), coverage of nominal 95% confidence intervals, and false discovery rates. The simulations were designed to assess estimator behavior in settings where selection depended on observed variables and the weighting model had access to the selection-related covariate.

Figure 6 compares bias, MSE, coverage, and FDR when estimating *β*_1_(*v*) using weighted and unweighted methods. Under the correctly specified model, weighted and unweighted regression models performed similarly. Both approaches produced approximately unbiased estimates and small MSE values, with average MSE values across vertices of 8.07 *×* 10^−5^ for the unweighted method and 9.37 *×* 10^−5^ for the weighted method. Both methods achieved coverage rates close to the nominal 0.95 level and similar FDR values for most vertices.

**Figure 6:**
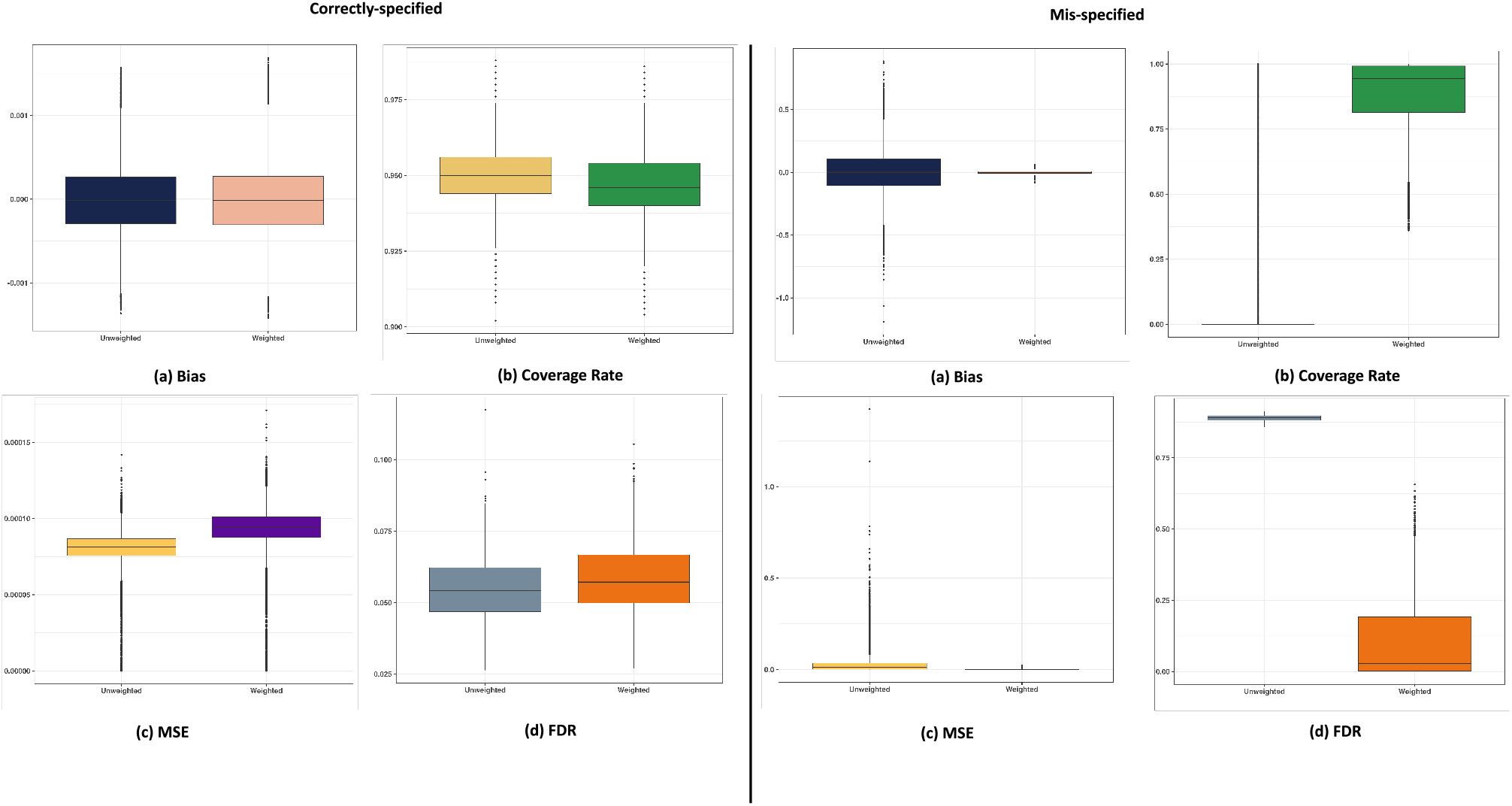
Simulation: Comparison of bias, mean squared error (MSE), coverage rates of 95% confidence intervals, and false discovery rates when estimating *β*_1_(*v*) using weighted and unweighted methods with both correctly specified and misspecified models.

Under the misspecified model, in which the outcome model omitted the interaction between the exposure and the selection-related covariate, weighted and unweighted estimates differed substantially. The unweighted method showed larger bias and greater variability, with vertex-wise biases ranging from −1.2 to 0.88. In contrast, the weighted method had smaller biases, ranging from −0.08 to 0.06. The average MSE across vertices was 0.026 for the unweighted model and 4.9 *×* 10^−4^ for the weighted model. The weighted method also showed improved coverage and lower FDR relative to the unweighted method in this simulation setting.

These results demonstrate that, when selection depends on observed variables and the target estimand is a population-average coefficient function, weighting can improve estimation under certain forms of model misspecification. However, the simulation represents a controlled setting and does not capture all complexities of ABCD imaging-data availability and quality-control processes. In particular, the simulations do not establish that weighting fully addresses selection processes driven by unmeasured variables.

## 4 Discussion

This study developed and evaluated a population-weighted image-on-scalar regression framework for analyzing high-dimensional neuroimaging outcomes when the analytic imaging sample may differ from the intended target population. The central methodological question was whether incorporating population and imaging-subsample weights changes estimated spatial associations between cognitive performance and working-memory-related fMRI activation. Across simulations and the ABCD application, our results show that weighting can affect estimated coefficient maps, statistical detection, and predictive performance, and that the magnitude and direction of these effects depend on model specification. In particular, differences between weighted and unweighted analyses were more pronounced in models without covariate adjustment and were attenuated, but not eliminated, after adjustment for measured covariates. These findings highlight the importance of clearly defining the target population, estimand, weighting assumptions, and covariate-adjustment strategy when using large-scale neuroimaging data for population-level inference.

Our ABCD application should be interpreted primarily as a methodological demonstration of how weighting can be incorporated into image-on-scalar regression, rather than as a definitive substantive analysis of working-memory activation in the ABCD Study. The empirical analysis used ABCD Curated Annual Release 2.0.1, and later releases include updated n-back contrasts. Therefore, the substantive neuroimaging findings reported here should be interpreted cautiously. Nevertheless, the application provides a useful case study because it illustrates how estimated brain–cognition association maps can differ when analyses target the available imaging analytic sample versus a population-average estimand represented through population and imaging-subsample weights.

In the ABCD application, both weighted and unweighted image-on-scalar regression models identified spatially distributed associations between the *g*-factor and the 2-back versus 0-back working-memory contrast. Regardless of whether sample weights were applied, our vertex detection results showed that specific brain regions, primarily frontal, parietal, and cingulate regions, had a higher proportion of detected vertices with significant associations than other regions. These vertices were components of several broad-scale functional networks, including the FPN and DAN. We observed a positive association between the *g*-factor and the 2-back versus 0-back contrast, indicating that adolescents with higher out-of-scanner general cognitive ability exhibited increased brain activity during high versus low memory load tasks in regions overlapping with the FPN and DAN. These findings are consistent with previous research (Finn et al., 2017; Rosenberg et al., 2020; Menon and D’Esposito, 2022). Conversely, we also observed negative associations within the DMN, CON, and PON, consistent with previous studies demonstrating an ‘anticorrelation’ between these networks in healthy adolescents, as evidenced by other analyses using ABCD data (Owens et al., 2020; Daamen et al., 2015).

In addition to the general findings on the association between working memory and cognitive ability among adolescents, our data analysis revealed the impact of applying sample weights in image-on-scalar regression. Notably, the unweighted regression method tended to identify more vertices with significant associations compared to the weighted method, particularly in association cortices. There are two potential interpretations for regions detected exclusively by unweighted regression but not by weighted regression: 1) Sample-specific significance: These regions, including the insula, middle temporal gyrus, and cingulate cortex, may be related to the working memory task but are only significant within the ABCD imaging subsample. They are not significant once sample weights are incorporated, and therefore, they are not likely generalizable to the broader population. 2) Potential false discoveries: Some regions might be identified due to potential false discoveries. For instance, the primary motor region, primary somatosensory-hand cortex, and primary auditory cortex are not typically associated with working memory and cognitive tasks. On the other hand, the inferior temporal gyrus was detected only by the weighted image-on-scalar regression model and not by the unweighted model. Since the inferior temporal gyrus is commonly associated with working memory and cognitive ability according to Fiebach et al. (2007), this result suggests that incorporating sample weights in image-on-scalar regression may help identify brain regions with associations that are significant in larger populations but may be undetected when analyzing subsamples.

The influence of weighting also depended on covariate adjustment. In several regions, differences between weighted and unweighted detection were reduced after adjustment for measured control variables. This pattern is expected because covariate adjustment and weighting can both account, in different ways, for measured differences between the analytic sample and the target population. However, differences persisted in some regions, including portions of the lateral occipital and cingulate cortex. These persistent differences may reflect association heterogeneity, incomplete model specification, residual differences in the distribution of measured variables, or unmeasured factors related to imaging availability and brain activation. This observation is consistent with prior work identifying heterogeneous image-on-scalar associations in the ABCD Study, with latent subgroups differing in sociodemographic profiles and in patterns of association between the *g*-factor and working-memory activation (Lin et al., 2024). More generally, these results illustrate that including measured covariates as main effects does not necessarily make weighted and unweighted regression estimates equivalent, particularly when associations vary across subgroups or when relevant interactions are omitted.

Simulation studies reinforced the necessity of sample weighting, particularly when models are misspecified and do not include all relevant interactions. This is crucial because, in real-world neuroimaging studies, it is often impractical to include all potential interaction terms to account for all possible confounders. Under correctly specified models, weighted and unweighted estimators performed similarly in terms of bias, mean squared error, and interval coverage. Under model misspecification, particularly when relevant interactions were omitted, weighting improved estimation of the target population-average coefficient function in the simulation settings considered. Thus, sample weights are particularly critical in analytic contexts where the model structure may not fully reflect the underlying data. In conjunction with our real-world analyses using the ABCD data, we observed substantial differences when utilizing weighted methods. Nevertheless, the simulations demonstrate that the proposed weighted estimation procedure behaves as expected under measured-variable assumptions, but they do not establish that weighting fully addresses the more complex imaging-selection and quality-control processes present in ABCD data.

However, several limitations must be acknowledged. First, as discussed above, the ABCD application used Curated Annual Release 2.0.1. Later ABCD releases contain important updates to n-back contrasts. We therefore interpret the ABCD results as a methodological application rather than as definitive substantive findings about working-memory activation. Future analyses should repeat the application using the most recent release.

Second, weighting can only address differences captured by measured variables included in the weighting procedure. The imaging-subsample weights were constructed using baseline variables available for both participants with and without usable fMRI data. Some potentially important drivers of imaging availability, such as detailed motion metrics, task compliance measures, scanner/protocol features, or processing-specific quality-control indicators, may be associated with brain activity, cognitive performance, or heterogeneity in brain–cognition associations, are omitted in the adjustment, so the weighted estimates may still differ from the desired population-average estimand. Thus, our analysis evaluates the influence of weighting under measured-variable assumptions and should not be interpreted as definitively eliminating imaging-selection bias.

Third, the ABCD baseline weights primarily align the baseline cohort with external sociodemographic benchmarks. They may not fully address all characteristics related to recruitment into the ABCD Study, participation in imaging, or brain–cognition associations. The weighted estimates should therefore be interpreted as estimates that incorporate available population and subsample weighting information, rather than as definitive estimates of population structure or population-level neurobiology (Wade et al., 2026).

Fourth, the impact of weighting is estimand- and model-dependent. The present analysis focused on the association between the *g*-factor and working-memory-related fMRI activation. Weighting may have different consequences for other exposures, outcomes, task contrasts, developmental periods, or covariate specifications. Similarly, population weighting is aimed at estimating overall population-average associations; it does not by itself ensure reliable inference for small subgroups. Subgroup-specific estimands may require additional modeling strategies, sufficient subgroup sample sizes, and possibly subgroup-specific weighting or hierarchical modeling approaches (Si, 2025; Si et al., 2020; Gelman et al., 2026). Fifth, our inference procedures treat the constructed weights as fixed. This is common in many applied weighted analyses, but it may understate uncertainty because it does not fully propagate uncertainty from estimating imaging-subsample propensity weights.

Finally, our analysis focuses on association estimates and external validity rather than causal effects. Weighting for population representation addresses one component of generalizability, but causal interpretation would additionally require assumptions and methods for internal validity, including careful control of confounding between the exposure and outcome. In the present study, the *g*-factor was treated as an exposure or predictor of interest in an associational regression framework. Therefore, our findings should be interpreted as weighted and unweighted associations, not as causal effects of cognitive ability on brain activation.

Several directions for future research follow from this work. First, the current model was applied to cortical surface fMRI data; extending the framework to jointly accommodate cortical and subcortical regions would be valuable, particularly for tasks involving reward, emotion, or motivational circuitry. Second, future work should examine alternative cognitive and behavioral measures beyond the *g*-factor and evaluate whether weighting has similar consequences across different predictors and task paradigms. Third, the proposed framework should be applied to recent ABCD data releases when accessible (as discussed above) and other large-scale cohorts, such as the UK Biobank, where population structure, sampling design, and weighting strategies differ. Finally, open-source software and computational improvements are needed to make weighted image-on-scalar regression more accessible, scalable, and reproducible for population neuroscience applications.

In summary, this study shows that incorporating population and imaging-subsample weights into image-on-scalar regression can change estimated brain–cognition association maps and statistical conclusions, particularly when model specification is limited or when the analytic imaging sample differs from the target population on measured characteristics. These findings support the use of weighting as a sensitivity and estimation tool for population-average neuroimaging analyses, while also underscoring that weighted estimates must be interpreted in light of the target estimand, available covariates, positivity, data quality, and the assumptions required for population inference.

## Author Contributions

YS: conceptualization, supervision, project administration, resources, methodology, formal analysis, validation, writing—original draft, writing—review and editing. ZL: conceptualization, software, methodology, validation, data curation, formal analysis, writing—original draft. JK: conceptualization, supervision, resources, methodology, formal analysis, validation, writing—review and editing. MFM: writing—review and editing. CS: data curation, writing—review and editing.

## Acknowledgments

The work was supported by the National Institutes of Health (NIH) with grants: R21HD105204, U01MD017867 and T32AA007477.

The Adolescent Brain Cognitive Development (ABCD) Study (abcdstudy.org) is a multisite, longitudinal study designed to recruit more than 10,000 children age 9 to 10 and follow them over 10 years into early adulthood. The ABCD Study is supported by the NIH and additional federal partners under award numbers U01DA041022, U01DA041028, U01DA041048, U01DA041089, U01DA041106, U01DA041117, U01DA041120, U01DA041134, U01DA041148, U01DA041156, U01DA041174, U24DA041123, and U24DA041147. A full list of supporters is available at abcdstudy.org/nih-collaborators. A listing of participating sites and a complete listing of the study investigators can be found at abcdstudy.org/principal-investigators.html. ABCD consortium investigators designed and implemented the study and/or provided data but did not necessarily participate in analysis or writing of this report. This manuscript reflects the views of the authors and may not reflect the opinions or views of the NIH or ABCD consortium investigators.

## Data availability

The ABCD data used in this paper can be obtained from the NIMH Data Archive (NDA) under Collection 2573 with a data use agreement. The code is available at GitHub: https://github.com/zikaiLin/weighted_image_on_scalar.

## References

Akshoomoff, N., J. L. Beaumont, P. J. Bauer, S. S. Dikmen, R. C. Gershon, D. Mungas, J. Slotkin, D. Tulsky, S. Weintraub, P. D. Zelazo, et al. (2013). VIII. NIH toolbox cognition battery (CB): Composite scores of crystallized, fluid, and overall cognition. Monographs of the Society for Research in Child Development 78 (4), 119–132.

Binder, D. (1983). On the variances of asymptotically normal estimators from complex surveys. International Statistical Review 51 (3), 279–292.

Casey, B., T. Cannonier, M. I. Conley, A. O. Cohen, D. M. Barch, M. M. Heitzeg, M. E. Soules, T. Teslovich, D. V. Dellarco, H. Garavan, et al. (2018). The Adolescent Brain Cognitive Development (ABCD) study: Imaging acquisition across 21 sites. Developmental Cognitive Neuroscience 32, 43–54.

Christophel, T. B., P. C. Klink, B. Spitzer, P. R. Roelfsema, and J.-D. Haynes (2017). The distributed nature of working memory. Trends in Cognitive Sciences 21 (2), 111–124.

Daamen, M., J. G. Bäuml, L. Scheef, C. Sorg, B. Busch, N. Baumann, P. Bartmann, D. Wolke, A. Wohlschläger, and H. Boecker (2015). Working memory in preterm-born adults: Load-dependent compensatory activity of the posterior default mode network. Human Brain Mapping 36 (3), 1121–1137.

Dick, A. S., S. Hawes, M. Bayat, M. Curtis, D. A. Lopez, P. Parekh, D. Pecheva, R. Gonzalez, S. G. Heeringa, J. Linkersdörfer, R. Loughnan, L. Aguinaldo, K. J. Sher, T. E. Nichols, S. F. Tapert, M. Neale, A. Dale, and W. K. Thompson (2026). The Adolescent Brain Cognitive Development (ABCD) Study: Ten years of statistical and methodological contributions. Developmental Cognitive Neuroscience 81, 101779.

Diebolt, J. and E. H. Ip (1996). Stochastic EM: Method and application. In Markov chain Monte Carlo in practice, pp. 259–273. Springer.

Dotson, V. M. and A. Duarte (2020). The importance of diversity in cognitive neuroscience. Annals of the New York Academy of Sciences 1464 (1), 181–191.

Falk, E. B., L. W. Hyde, C. Mitchell, J. Faul, R. Gonzalez, M. M. Heitzeg, D. P. Keating, K. M. Langa, M. E. Martz, J. Maslowsky, et al. (2013). What is a representative brain? Neuroscience meets population science. Proceedings of the National Academy of Sciences 110 (44), 17615–17622.

Fiebach, C., A. Friederici, E. Smith, and D. Swinney (2007). Lateral inferotemporal cortex maintains conceptual—semantic representations in verbal working memory. Journal of Cognitive Neuroscience 19 (12), 2035–2049.

Finn, A. S., J. E. Minas, J. A. Leonard, A. P. Mackey, J. Salvatore, C. Goetz, M. R. West, C. F. Gabrieli, and J. D. Gabrieli (2017). Functional brain organization of working memory in adolescents varies in relation to family income and academic achievement. Developmental Science 20 (5), e12450.

Garavan, H., H. Bartsch, K. Conway, A. Decastro, R. Goldstein, S. Heeringa, T. Jernigan, A. Potter, W. Thompson, and D. Zahs (2018). Recruiting the ABCD sample: Design considerations and procedures. Developmental Cognitive Neuroscience 32, 16–22.

Gard, A. M., L. W. Hyde, S. G. Heeringa, B. T. West, and C. Mitchell (2023). Why weight? Analytic approaches for large-scale population neuroscience data. Developmental Cognitive Neuroscience 59, 101196.

Gelman, A., Y. Si, B. T. West, and J. Gabry (2026). MRPW: Regression, poststratification, and small-area estimation with sampling weights. https://sites.stat.columbia.edu/gelman/research/unpublished/weightregression.pdf.

Ghosal, S. and A. Van der Vaart (2017). Fundamentals of nonparametric Bayesian inference, Volume 44. Cambridge University Press.

Gordon, E., T. Laumann, B. Adeyemo, J. Huckins, W. Kelley, and S. Petersen (2016). Generation and evaluation of a cortical area parcellation from resting-state correlations. Cerebral cortex 26 (1), 288–303.

Gray, J. R., C. F. Chabris, and T. S. Braver (2003). Neural mechanisms of general fluid intelligence. Nature neuroscience 6 (3), 316–322.

Hagler Jr, D. J., S. Hatton, M. D. Cornejo, C. Makowski, D. A. Fair, A. S. Dick, M. T. Sutherland, B. Casey, D. M. Barch, M. P. Harms, et al. (2019). Image processing and analysis methods for the Adolescent Brain Cognitive Development Study. Neuroimage 202, 116091.

Han, Z., G. Yang, T. Liu, S. Funahashi, X.-N. Zuo, and T. Yan (2025). Enriching population diversity in neuroscience. Science Bulletin 70 (16), 2560–2564.

Hawes, S. W., A. K. Littlefield, D. A. Lopez, K. J. Sher, E. L. Thompson, R. Gonzalez, L. Aguinaldo, A. R. Adams, M. Bayat, A. L. Byrd, L. F. C. de Araujo, A. Dick, S. F. Heeringa, C. M. Kaiver, S. M. Lehman, L. Li, J. Linkersdörfer, T. J. Maullin-Sapey, M. C. Neale, T. E. Nichols, S. Perlstein, S. F. Tapert, C. E. Vize, M. Wagner, R. Waller, and W. K. Thompson (2025). Longitudinal analysis of the ABCD® study. Developmental Cognitive Neuroscience 72, 101518.

Heeringa, S. G. and P. A. Berglund (2020). A guide for population-based analysis of the Adolescent Brain Cognitive Development (ABCD) Study baseline data. BioRxiv, 2020–02.

LeWinn, K., M. Sheridan, K. Keyes, A. Hamilton, and K. McLaughlin (2017). Sample composition alters associations between age and brain structure. Nat Communication 8 (1), 874.

Li, L., M. Bayat, T. B. Hayes, W. K. Thompson, A. M. Gard, and A. Dick (2025). Missing data approaches for longitudinal neuroimaging research: Examples from the adolescent brain and cognitive development (abcd) study. Developmental Cognitive Neuroscience 74.

Lin, Z., Y. Si, and J. Kang (2024). Latent subgroup identification in image-on-scalar regression. Annals of Applied Statistics 18 (1), 468–486.

Menon, V. and M. D’Esposito (2022). The role of PFC networks in cognitive control and executive function. Neuropsychopharmacology 47 (1), 90–103.

Owens, M. M., D. Yuan, S. Hahn, M. Albaugh, N. Allgaier, B. Chaarani, A. Potter, and H. Garavan (2020). Investigation of psychiatric and neuropsychological correlates of default mode network and dorsal attention network anticorrelation in children. Cerebral Cortex 30 (12), 6083–6096.

Rosenberg, M. D., S. A. Martinez, K. M. Rapuano, M. I. Conley, A. O. Cohen, M. D. Cornejo, D. J. Hagler, W. J. Meredith, K. M. Anderson, T. D. Wager, et al. (2020). Behavioral and neural signatures of working memory in childhood. Journal of Neuroscience 40 (26), 5090–5104.

Rudolph, J., Y. Zhong, P. Duggal, S. Mehta, and L. B (2023). Defining representativeness of study samples in medical and population health research. BMJ Medicine 2 (1), e000399.

Rust, K. (1985). Variance estimation for complex estimators in sample surveys. Journal of Official Statistics 1 (4), 381–397.

Si, Y. (2025). On the use of auxiliary variables in multilevel regression and poststratification. Statistical Science 40 (2), 272–288.

Si, Y., S. Lee, and S. G. Heeringa (2024). Population weighting in statistical analysis. JAMA Internal Medicine 184 (1), 98–99.

Si, Y., R. J. Little, Y. Mo, and N. Sedransk (2023). A case study of nonresponse bias analysis in educational assessment surveys. Journal of Educational and Behavioral Statistics 48 (3), 271–295.

Si, Y., R. J. Little, Y. Mo, and N. Sedransk (2024). Nonresponse bias analysis in longitudinal studies: A comparative review with an application to the Early Childhood Longitudinal Study. International Statistical Review 92 (3), 383–405.

Si, Y., R. Trangucci, J. S. Gabry, and A. Gelman (2020). Bayesian hierarchical weighting adjustment and survey inference. Survey Methodology 46 (2), 181–214.

Si, Y., B. T. West, P. Veliz, M. E. Patrick, J. E. Schulenberg, D. D. Kloska, Y. M. Terry-McElrath, and S. E. McCabe (2022). An empirical evaluation of alternative approaches to adjusting for attrition when analyzing longitudinal survey data on young adults’ substance use trajectories. International Journal of Methods in Psychiatric Research 31 (3), e1916.

Sternberg, R. J. and E. L. Grigorenko (2002). The general factor of intelligence: How general is it? Psychology Press.

Thompson, W. K., D. M. Barch, J. M. Bjork, R. Gonzalez, B. J. Nagel, S. J. Nixon, and M. Luciana (2019). The structure of cognition in 9 and 10 year-old children and associations with problem behaviors: Findings from the ABCD study’s baseline neurocognitive battery. Developmental Cognitive Neuroscience 36, 100606.

Wade, N. E., Y. Si, S. F. Tapert, J. Linkersdörfer, K. M. Lisdahl, H. R. Moore, L. Tally, B. Das, M. A. Huestis, A. L. Wallace, R. M. Sullivan, V. Szpak, L. Zhang, L. Ziemer, and W. K. Thompson (2026). Weighted prevalence of biochemically verified substance use in healthy adolescents across the usa. Addiction.

Waiter, G. D., I. J. Deary, R. T. Staff, A. D. Murray, H. C. Fox, J. M. Starr, and L. J. Whalley (2009). Exploring possible neural mechanisms of intelligence differences using processing speed and working memory tasks: An fMRI study. Intelligence 37 (2), 199–206.

West, B. T., Y. Si, Y. Hu, S. E. McCabe, and P. V. and (2025). The role of weighting adjustment for attrition in longitudinal trajectory modeling: a simulation study. Communications in Statistics - Simulation and Computation 54 (3), 866–888.

White, H. (1980). A heteroskedasticity-consistent covariance matrix estimator and a direct test for heteroskedasticity. Econometrica: journal of the Econometric Society 48 (4), 817–838.

Williams, C. K. and C. E. Rasmussen (2006). Gaussian processes for machine learning. MIT press Cambridge, MA.

Yu, S., G. Wang, L. Wang, and L. Yang (2021). Multivariate spline estimation and inference for image-on-scalar regression. Statistica Sinica 31 (3), 1463–1487.

Zeng, Z., M. Li, and M. Vannucci (2022). Bayesian image-on-scalar regression with a spatial global-local spike-and-slab prior. Bayesian Analysis 1 (1), 1–26.

Zhu, H., J. Fan, and L. Kong (2014). Spatially varying coefficient model for neuroimaging data with jump discontinuities. Journal of the American Statistical Association 109 (507), 1084–1098.

